# Application of respiratory metagenomics for COVID-19 patients on the intensive care unit to inform appropriate initial antimicrobial treatment and rapid detection of nosocomial transmission

**DOI:** 10.1101/2020.11.26.20229989

**Authors:** Themoula Charalampous, Adela Alcolea-Medina, Luke B. Snell, Tom G.S Williams, Rahul Batra, Luigi Camporota, Christopher I.S. Meadows, Duncan Wyncoll, Nicholas A. Barrett, Carolyn J. Hemsley, Lisa Bryan, William Newsholme, Sara E. Boyd, Anna Green, Ula Mahadeva, Amita Patel, Penelope R. Cliff, Andrew J. Page, Justin O’Grady, Jonathan D. Edgeworth

## Abstract

**Background:** Clinical metagenomics (CMg) is being evaluated for translation from a research tool into routine diagnostic service, but its potential to significantly improve management of acutely unwell patients has not been demonstrated. The SARS-CoV-2 pandemic provides impetus to determine that benefit given increased risk of secondary infection and nosocomial transmission by multi-drug resistant (MDR) pathogens linked with expansion of critical care capacity.

**Methods:** Prospective evaluation of CMg using nanopore sequencing was performed on 43 respiratory samples over 14 weeks from a cohort of 274 intubated patients across seven COVID-19 intensive care units.

**Results:** Bacteria or fungi were cultured from 200 (73%) patients, with a predominance of *Klebsiella spp*. (31%) and *C. striatum* (7%) amongst other common respiratory pathogens. An 8 hour CMg workflow was 93% sensitive and 81% specific for bacterial identification compared to culture, and reported presence or absence of β-lactam resistance genes carried by *Enterobacterales* that would modify initial guideline-recommended antibiotics in every case. CMg was also 100% concordant with quantitative PCR for detecting *Aspergillus fumigatus* (4 positive and 39 negative samples). Single nucleotide polymorphism (SNP)-typing using 24 hour sequence data identified an MDR-*K. pneumoniae* ST307 outbreak involving 4 patients and an MDR-*C. striatum* outbreak potentially involving 14 patients across three ICUs.

**Conclusion:** CMg testing for ICU patients provides same-day pathogen detection and antibiotic resistance prediction that significantly improves initial treatment of nosocomial pneumonia and rapidly detects unsuspected outbreaks of MDR-pathogens.

## Introduction

The intensive care unit (ICU) is a dynamic environment with frequent staff-contact for invasive monitoring, interventions and personal care that together introduce risk of secondary or nosocomial infection (1). Invasive ventilation can introduce organisms into the lungs causing ventilator-acquired pneumonia (VAP) which carries high attributable mortality and drives up to 70% of antimicrobial prescribing (2). Typically, patients with suspected VAP receive guideline-directed empiric antibiotics until culture results return, typically 2-4 days later, although therapy isn’t always adjusted when results are returned (3, 4). Invasive pulmonary aspergillosis (IPA) is also increasingly recognised on ICU particularly with severe influenza (5, 6) and after host immunosuppression, but culture lacks sensitivity, biomarker tests have low specificity and long turnaround times (7) and gold-standard histopathology is rarely used (8).

SARS-CoV-2 has put considerable strain on ICUs, due to expansion of bed capacity, which has potential to increase nosocomial infection and antimicrobial resistance (AMR). High prevalence of Gram-negative bacteria (GNB) particularly *Klebsiella* spp. have been reported (9-11) and there are reports of secondary IPA (12-15). COVID-19 patients also receive steroid therapy, which could exacerbate bacterial or fungal infection (16, 17). The COVID-19 pandemic therefore re-enforces the need for rapid comprehensive diagnostics to improve empiric therapy and help prevent emergence and transmission of MDR-organisms.

Clinical metagenomic (CMg) using nanopore technology has potential to meet these needs due to its unbiased pan-microbial coverage (18, 19) and ability to provide real-time data acquisition and analysis (20). It has been evaluated for respiratory, urinary tract and prosthetic joint infections (20-23), however, the full clinical potential required for laboratories and clinical teams to change their long-standing practice has not been demonstrated. We therefore prospectively assessed whether CMg testing of respiratory samples from COVID-19 patients with suspected secondary bacterial or fungal pneumonia, could significantly improve their initial antimicrobial treatment and detect outbreaks affecting a large COVID-19 patient cohort across 7 ICUs.

## Results

### Clinical and microbiological characteristics of COVID-19 patients

274 consecutive invasively-ventilated COVID-19 patients were admitted between March 20^th^ and June 24^th^ 2020 (Table 1), including 103 inter-hospital transfers for specialist care and assessment for extracorporeal membrane oxygenation (ECMO). Median age was 56 and 71% were male. Median length of hospital-stay was 19 days (Interquartile range (IQR) 12-37) and 196 patients (71%) were discharged alive from hospital. 763 respiratory samples were processed from 225 (82%) patients, with organisms isolated from at least one sample in 77% of patients. The main GNB were *Klebsiella spp*. (31%) *Citrobacter spp*. (23%), *E. coli* (7%), and *P. aeruginosa* (7%). The main Gram-positive bacteria were *S. aureus* (10%), *Enterococcus spp*. (7%) and *C. striatum* (7%). *C. albicans*, other *Candida spp*. and *Aspergillus spp*. were cultured from 28%, 8% and 2% of patients respectively. CMg was performed on 43 samples from 34 patients selected because of strong clinical suspicion of secondary infection. All respiratory pathogens cultured from the whole cohort were represented in CMg samples including 5 of the 6 patients from which *Aspergillus spp*. was isolated.

**Table 1.** Clinical characteristics and results of routine microbiological tests performed on intubated COVID-19 patients across 7 linked dedicated COVID-19 intensive care units on Guy’s and St Thomas’ hospital sites

|  | All<br>(n = 274) | Metagenomics<br>group<sup>a</sup><br>(n = 34) | Non-metagenomics<br>group<br>(n = 240) |
| --- | --- | --- | --- |
| Median age (IQR) | 56 (45-63) | 52 (41-58) | 56 (46-63) |
| Sex – Male | 195 (71%) | 23 (70%) | 172 (72%) |
| Ethnicity |  |  |  |
| White | 98 (36%) | 15 (45%) | 83 (35%) |
| Black and Minority Ethnicities | 110 (40%) | 15 (45%) | 95 (41%) |
| Not known | 65 (24%) | 3 (9%) | 62 (26%) |
| Mortality | 78 (29%) | 8 (24%) | 71 (30%) |
| Length of stay (IQR) | 19 days (12-37) | 32 days (24-47) | 17 days (11-32) |
| **Respiratory Cultures in ITU<sup>b</sup>** |  |  |  |
| Median samples per patient (IQR) | 2 (1 – 4) | 4.5 (3 – 6) | 2 (1 – 3) |
| Total number of samples / Patients tested | 763 / 225 | 183 / 33 | 580 / 192 |
| **Organisms from respiratory culture whilst in ICU (Number of individuals who ever had the following organisms in any sample)** |  |  |  |
| *Klebsiella spp.* | 86 (31%) | 19 (56%) | 67 (28%) |
| *Staphylococcus aureus* | 27 (10%) | 3 (9%) | 24 (10%) |
| *Citrobacter spp.* | 23 (8%) | 6 (18%) | 17 (7%) |
| *Escherichia coli* | 19 (7%) | 4 (12%) | 15 (6%) |
| *Pseudomonas spp.* | 19 (7%) | 2 (6%) | 17 (7%) |
| *Corynebacterium striatum* | 18 (7%) | 8 (24%) | 10 (4%) |
| *Enterococcus spp.* | 18 (7%) | 4 (12%) | 14 (6%) |
| *Serratia spp.* | 14 (5%) | 2 (6%) | 12 (5%) |
| *Enterobacter spp.* | 10 (4%) | 1 (3%) | 9 (4%) |
| *Haemophilus spp.* | 8 (3%) | 2 (6%) | 6 (3%) |
| *Stenotrophomonas maltophilia* | 7 (3%) | 2 (6%) | 5 (2%) |
| *Proteus spp.* | 4 (1%) | 4 (12%) | 0 (0%) |
| *Morganella spp.* | 3 (11%) | 1 (3%) | 2 (1%) |
| *Moraxella spp.* | 2 (1%) | 0 | 2 (1%) |
| *Acinetobacter spp.* | 2 (1%) | 1 (3%) | 1 (0%) |
| *Streptococcus pyogenes* | 1 (0.3%) | 0 | 1 (0%) |
| *Candida albicans* | 76 (28%) | 12 (35%) | 64 (27%) |
| *Candida spp. (non albicans)* | 21 (8%) | 6 (18%) | 15 (6%) |
| *Aspergillus spp.* | 6 (2%) | 5 (15%) | 1 (0%) |
| No organisms isolated | 74 (27%) | 1 (3%) | 73 (30%) |
| **All clinically significant organisms from blood culture whilst in ICU (Number of individuals who ever had the following organisms in any sample)** |  |  |  |
| *Staphylococcus (non aureus)* | 36 (13%) | 7 (21%) | 29 (12%) |
| *Klebsiella spp.* | 19 (7%) | 4 (12%) | 15 (6%) |
| *Enterococcus spp.* | 8 (3%) | 1 (3%) | 7 (3%) |
| *Escherichia coli* | 5 (2%) | 0 (0%) | 5 (2%) |
| *Pseudomonas spp.* | 4 (1%) | 0 (0%) | 4 (2%) |
| *Staphylococcus aureus* | 2 (1%) | 1 (3%) | 1 (0%) |
| *Stenotrophomonas maltophilia* | 2 (1%) | 1 (3%) | 1 (0%) |
| *Citrobacter spp.* | 1 (0%) | 0 (0%) | 1 (0%) |
| *Enterobacter spp.* | 1 (0%) | 0 (0%) | 1 (0%) |
| *Proteus spp.* | 1 (0%) | 0 (0%) | 1 (0%) |
| *Candida (non albicans)* | 2 (1%) | 0 (0%) | 2 (1%) |
| *Candida albicans* | 0 (0%) | 0 (0%) | 0 (0%) |
| Contaminants only<sup>c</sup> | 12 (44%) | 4 (12%) | 8 (3%) |
| No organisms isolated | 196 (72%) | 16 (47%) | 180 (75%) |
| **Galactomannans (GAL)** |  |  |  |
| **Bronchoalveolar lavage (BAL) GALs** |  |  |  |
| Number of tests / Patients tested | 76 / 51 | 28 / 18 | 48 / 33 |
| Positive tests / Patients positive | 10 / 8 | 9 / 7 | 1 / 1 |
| **Serum GALs** |  |  |  |
| Number of tests / Patients tested | 119 / 74 | 35 / 22 | 84 / 52 |
| Positive tests / Patients positive | 7 / 7 | 3 / 3 | 4 / 4 |

<sup>a</sup>1 patient was SARS-CoV-2 RNA PCR negative but had clinical diagnosis of COVID-19. <sup>b</sup>1 patient in the metagenomics group and 48 patients from the non-metagenomics group had no respiratory specimens collected while on ITU. <sup>c</sup>The significance of all blood-culture isolates was independently determined by the infectious diseases team as part of routine daily clinical consult service ward rounds on the ICUs

There were 79 clinically-significant blood stream infections (BSIs) of which 36 were coagulase-negative *Staphylococci. Klebsiella spp*. were the second most frequent (n=19) representing 57% of all GNB-BSIs, comprising 12 *K. pneumoniae*, 5 *K. aerogenes*, 1 *K. oxytoca* and 1 *K. variicola* (Supplementary Table 1). Nine *K. pneumoniae* BSIs had acquired β-lactam resistance and, overall, 15 (79%) BSIs caused by *Klebsiella spp* and 70/94 (74%) patients with respiratory *Klebsiella spp*. isolates had phenotypic resistance to first-line empiric antibiotic treatment for nosocomial pneumonia (piperacillin-tazobactam). There were few BSIs with other organisms: *E. coli* (n=5), *P. aeruginosa* (n=4) and *S. aureus* (n=2).

### Performance of CMg compared with routine culture for pathogen detection

Potential respiratory pathogens were cultured from 27/43 (63%) samples tested by CMg and 16 samples (14 patients) were reported either as no growth or not clinically significant. CMg identified 25/27 culture-reported pathogens (93% sensitive; 95% CI, 76-99%) (Table 2). It did not report scanty *K. aerogenes* in two polymicrobial samples (S44 and S45) that were present below pre-defined thresholds (supplementary Table 2A). CMg reported 3 additional pathogens in 3 culture negative samples: 1 *A. fumigatus* and 2 *S. aureus* (specificity 81% (95%CI, 54-96%). The additional *A. fumigatus* was from a patient growing *A. fumigatus* in other respiratory samples (S55). One *S. aureus* false positive sample contained high levels of *S. epidermidis* (>15,000 reads) and a proportion of the sequence reads can be incorrectly identified as *S. aureus* (S16). The other additional *S. aureus* was in a sample (S41) containing multiple commensals, so may have been missed by culture-plate reading (suppl. Table 6).

**Table 2.** Comparison of pathogens reported by routine culture with metagenomics sequencing in respiratory samples

| Patient ID | Sample ID | ICU ward<sup>a</sup> | Semi-quantitative routine culture report<sup>b</sup> | Pathogens identified by metagenomic sequencing |
| --- | --- | --- | --- | --- |
| 26 | S35 | 6 | *Acinetobacter* spp. (L) | *A. baumannii* |
| 100 | S39 | 5 | *C.koseri* (H) | *C. koseri*<br>*K. pneumoniae* |
| 121 | S37 | 3 | *P.mirabilis* (M)<br>*M.morganii* (M) | *P. mirabilis*<br>*M. morgannii*<br>*K. pneumoniae* |
| 177 | S36 | 3 | *S. aureus* (H) | *S. aureus* |
| 196 | S42 | 3 | *B. cenocepacia* (L) | *Burkholderia* spp. |
| 400 | S49 | 5 | *K. pneumoniae* (M) | *K. pneumoniae* |
| 408 | S20 | 5 | *S. aureus* (M) | *S. aureus* |
| 441 | S21 | 3 | *E.cloacae* (M) | *E. cloacae* |
|  | S51 | 5 | *S. aureus* (L)<br>*C. koseri* (L) | *S. aureus*<br>*C. koseri* |
| 550 | S10 | 2 | *K. pneumoniae* (M) | *K. pneumoniae* |
| 563 | S28 | 4 | *Aspergillus* (S) | *A. fumigatus*<br>*S. aureus* |
| 613 | S18 | 4 | Negative | Negative |
| 618 | S45 | 5 | *C. striatum* (M)<br>*K. aerogenes* (S) | *C. striatum* |
| 677 | S52 | 7 | *K. aerogenes* (L) | *K. aerogenes*<br>*C. striatum* |
|  | S54 | 7 | *C. striatum* (M) | *C. striatum* |
|  | S63 | 4 | *K. pneumoniae* (M)<br>*C. striatum* (M) | *C. striatum*<br>*K. pneumoniae* |
| 727 | S53 | 7 | Negative | Negative |
| 740 | S30 | 3 | Negative | Negative |
|  | S59 | 3 | *K. pneumoniae* (M)<br>*C. striatum* (M) | *K. pneumoniae*<br>*C. striatum* |
| 749 | S40 | 5 | Negative | Negative |
|  | S62 | 3 | *K. aerogenes* (L)<br>*C. striatum* (L) | *K. aerogenes*<br>*C. striatum* |
| 815 | S25 | 5 | Negative | Negative |
|  | S46 | 5 | *C. koseri* (M) | *C. koseri* |
| 855 | S41 | 5 | Negative | *S. aureus* |
| 872 | S11 | 3 | *K. pneumoniae* (H) | *K. pneumoniae* |
|  | S61 | 3 | *P. mirabilis* (H)<br>*K. pneumoniae* (M) | *P. mirabilis*<br>*K. pneumoniae*<br>*C. koseri* |
| 1033 | S8 | 1 | *A. fumigatus* (S) | *A. fumigatus*<br>*K. oxytoca* |
| 1036 | S5 | 1 | Negative | Negative |
| 1054 | S31 | 3 | *K. pneumoniae* (L) | *K. pneumoniae* |
| 1065 | S16 | 1 | Negative | *S. aureus* |
|  | S19 | 1 | Negative | Negative |
| 1069 | S17 | 1 | *P. aeruginosa* (M) | *P. aeruginosa* |
| 1082 | S14 | 1 | Negative | Negative |
| 1092 | S27 | 1 | Negative | Negative |
| 1262 | S29 | 1 | Negative | Negative |
| 1292 | S44 | 1 | *S. marcescens* (L)<br>*K. aerogenes* (S) | *S. marcescens*<br>*C. freundii* |
| 1346 | S56 | 1 | *A. fumigatus* (S)<br>*P. mirabilis* (L) | *A. fumigatus*<br>*P. mirabilis* |
| 1440 | S33 | 1 | Negative | Negative |
| 1457 | S64 | 1 | Negative | Negative |
|  | S65 | 1 | Negative | Negative |
| 1503 | S1 | 1 | *K. aerogenes* (L) | *K. aerogenes* |
| 1512 | S34 | 1 | *K. pneumoniae* (L) | *K. pneumoniae* |
| 1538 | S55 | 3 | Negative | *A. fumigatus* |

<sup>a</sup>The seven dedicated adult COVID-19 ITUs comprise existing adult ITUs (1,3 and 4), a pre-purposed paediatric ITU (5), two post-operative recovery units (2 and 6) and a specialist chronic respiratory unit (7). <sup>b</sup>Reported growth by culture for each pathogen . H = heavy growth, M = moderate growth, L = light growth
Criteria for reporting organisms was $\geq 1\%$ of microbial classified reads and with a centrifuge score $\geq 2504$

CMg also identified 9 additional pathogens in 7 culture-positive samples (Table 2). 4 were identified by culture in other respiratory samples from those patients (*K. oxytoca* (S8), *K. pneumoniae* (S37), *C. striatum* (S52) and *C. koseri* (S61). All these additional CMg reported bacteria were considered true positives and clinically reportable.

### Impact of resistance gene detection on guideline-directed empiric antibiotic selection

Two-hour CMg sequencing data was analysed from 21 of 27 culture-positive samples where presence of acquired resistance could impact on guideline-directed treatment (Table 3). There was genotypic and phenotypic concordance in all but one sample. Extended spectrum β-lactamase (ESBL) genes were detected in 4 samples containing *Enterobacterales* with phenotypic resistance, *bla*_TEM_ genes in S31, S49 and S59, *bla*_SHV_ and *bla*_CTX-M_ in S31 and S59 and *bla*_SHV_ in S63. No β-lactamase resistance genes were found in 9 samples containing 10 susceptible *Enterobacterales* (Supplementary Table 3), but *bla*_TEM_ and *bla*_SHV_ genes were detected in a sample with *K. pneumoniae* having no reported phenotypic acquired resistance (S34). Resistance phenotypes could not be genotypically predicted in two samples with light bacterial-growth of *A. baumanni* (S35) and *K. aerogenes* (S62) due to low read count. No carbapenemases were detected in any sample and no SCC_mec_ elements in the two samples growing *S. aureus*. Genes conferring resistance against non-guideline recommended antibiotics were detected, all consistent with reported phenotypes (Table 3).

**Table 3.**
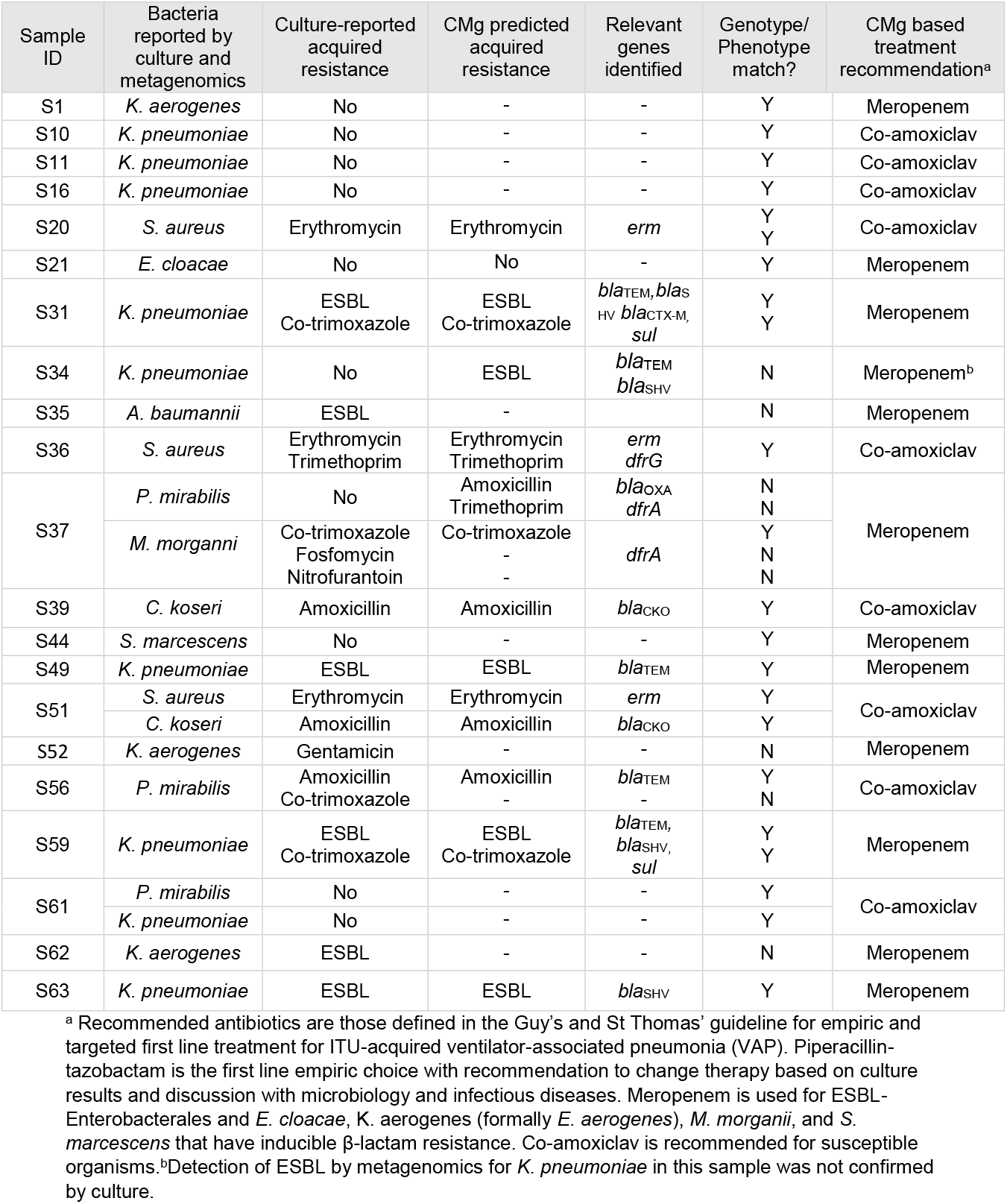
Comparison of CMg identified acquired genotypic resistance with phenotypic culture results and the impact on guideline-recommended antibiotic treatment

CMg data would have informed meropenem therapy in 11/21 cases, based on speciation in 7 (33%) and acquired resistance-genotype in 4 (19%). CMg data would have guided the use of co-amoxiclav therapy in 9 based on speciation combined with absence of β-lactamases (43%). In 1/21 (5%) cases CMg directed antibiotic choice was not consistent with culture - from S34 where identification of an ESBL was not phenotypically confirmed by culture.

### Comparison of methods for diagnosis of IPA

763 respiratory samples from 225 patients were cultured to identify *A. fumigatus*, and galactomannan (GM) antigen detection tests were requested on bronchoalveolar lavage and serum samples from 51(19%) and 74 (27%) patients, respectively. Nine patients had at least one mycology result consistent with IPA (Table 4). Four of five culture positive patients met original AspICU criteria (24) and all met modified AspICU criteria that do not require predisposing host factors (25). Four persistently culture negative patients had positive BAL-GM and met modified AspICU criteria (26); none of these had *Aspergillus* detected by CMg and qPCR.

**Table 4.** Mycological tests and clinical characteristics of patients with at least one result suggestive of invasive pulmonary Aspergillosis.

| Patient | *A. fumigatus* detection in respiratory samples |  |  |  | Galactomannan<br>(Positive/Tested) |  | AspICU – Putative Criteria (24) |  |  | ECMO |
| --- | --- | --- | --- | --- | --- | --- | --- | --- | --- | --- |
|  | Sample Number | CMg | qPCR (Cq) | Culture<br>(Positive/Tested) | BAL<br>> 1.0 | Serum<br>> 0.5 | Radiology | Clinical | Host |  |
| 563 | S28 | Positive | 31 | Positive |  |  | Yes | Yes | Yes – steroid | No |
|  | Other | ND | ND | 3 / 6 | 0 / 1 | 0 / 0 |  |  |  |  |
| 613 | S18 | Negative | >40 | Negative |  |  | Yes | Yes | No | No |
|  | Other | ND | ND | 2 / 2 | 1 / 1 | 0 / 0 |  |  |  |  |
| 677 | S63 | Negative | >40 | Negative |  |  | Yes | Yes | Yes - steroid | No |
|  | S54 | Negative | >40 | Negative |  |  |  |  |  |  |
|  | S52 | Negative | >40 | Negative |  |  |  |  |  |  |
|  | Other | ND | ND | 0 / 8 | 2 / 2 | 0 / 5 |  |  |  |  |
| 740 | S59 | Negative | >40 | Negative |  |  | Yes | Yes | Yes –<br>Leukaemia on<br>chemotherapy | No |
|  | S30 | Negative | >40 | Negative |  |  |  |  |  |  |
|  | Other | ND | ND | 0 / 16 | 1 / 4 | 1 / 2 |  |  |  |  |
| 1033 | S8 | Positive | 33 | Positive |  |  | Yes | Yes | Yes - steroid | Yes |
|  | Other | ND | ND | 0 / 0 | 1 / 1 | 1 / 1 |  |  |  |  |
| 1346 | S56 | Positive | 32 | Positive |  |  | Yes | Yes | Yes – steroid,<br>anakinra | Yes |
|  | Other | ND | ND | 0 / 3 | 1 / 1 | 0 / 2 |  |  |  |  |
| 1440 | S33 | Negative | >40 | Negative |  |  | Yes | Yes |  | Yes |
|  | Other | ND | ND | 0 / 4 | 1 / 2 | 0 / 1 |  |  | Yes – steroid,<br>anakinra |  |
| 1457 | S65 | Negative | >40 | Negative |  |  | Yes | Yes | Yes – steroid | Yes |
|  | S64 | Negative | >40 | Negative |  |  |  |  |  |  |
|  | Other | ND | ND | 0 / 9 | 2 / 2 | 0 / 2 |  |  |  |  |
| 1538 | S55 | Positive | 31 | Negative |  |  | Yes | Yes | Yes –<br>Lymphoma on<br>chemotherapy | No |
|  | Other | ND | ND | 4 / 5 | 0 / 0 | 1 / 1 |  |  |  |  |

ND = Not done

Two-hour CMg sequence data identified *A. fumigatus* reads in all of the 3 culture-positive samples that were tested by CMg (S8 [77 reads], S28 [2649 reads] and S56 [79 reads]) and a sample from a patient with *A. fumigatus* in other diagnostic samples (S55 [16 reads]) - see Supplementary table 2A. Probe-based qPCR (27) was 100% concordant with CMg (Table 4). One sample from a patient (S18) growing *A. fumigatus* in other samples was negative by culture, qPCR and metagenomic sequencing. There was discordance between GM tests and culture, and between BAL-GM and serum-GM. Four culture positive patients had at least one BAL-GM performed, with three patients having at least one positive BAL-GM result, and 2 of 3 culture-positive patients who were tested with serum-GM had a positive result.

Post-mortem histology from patient 563 with *A. fumigatus* identified by culture and CMg revealed a single 1cm x 1cm patch of IPA and no *A. fumigatus* in other organs. There was extensive diffuse alveolar damage and IPA was not reported to have contributed to death (Supplementary Fig.1).

### CMg detection of hospital transmission

Higher than anticipated prevalence of *Klebsiella spp*. and *C. striatum* raised the possibility of patient to patient transmission that was investigated by analysis of 24 hour CMg sequencing data.

### Klebsiella pneumoniae

Consensus sequence was generated using *K. pneumoniae* reads from 8 samples (8 patients). Different sequence-types (ST) were determined in four samples (S11, S34, S59 and S63). No ST could be determined for the remaining three samples (S10, S31 and S61), and S49 was excluded from analysis due to 3% genome coverage (Supplementary Table 5C). Comparison of high quality allele calls and pairwise comparison of bases from all samples showed S31 was similar to S59 (ST307) with 55 SNP-based differences from 4,892,921 bases (99.999% identical). This indicates a recent evolutionary history with differences likely due to nanopore sequencing errors. All other samples differed by tens of thousands of SNPs (Supplementary Table 5B).

Two additional patients (301 and 968) had a *K. pneumoniae*-BSI with identical broad resistance phenotype as CMg samples S31 and S59 (patient 1054 and 740 respectively). Pairwise comparison of SNP differences across all 4 genomes showed they were virtually identical with 5-55 SNP differences (see supplementary Table 5D). Epidemiological analysis found all 4 patients with this *K. pneumoniae* ST307 clone had overlapping stays on one ICU implicating an unsuspected outbreak (Fig. 1A).

**Figure 1.**
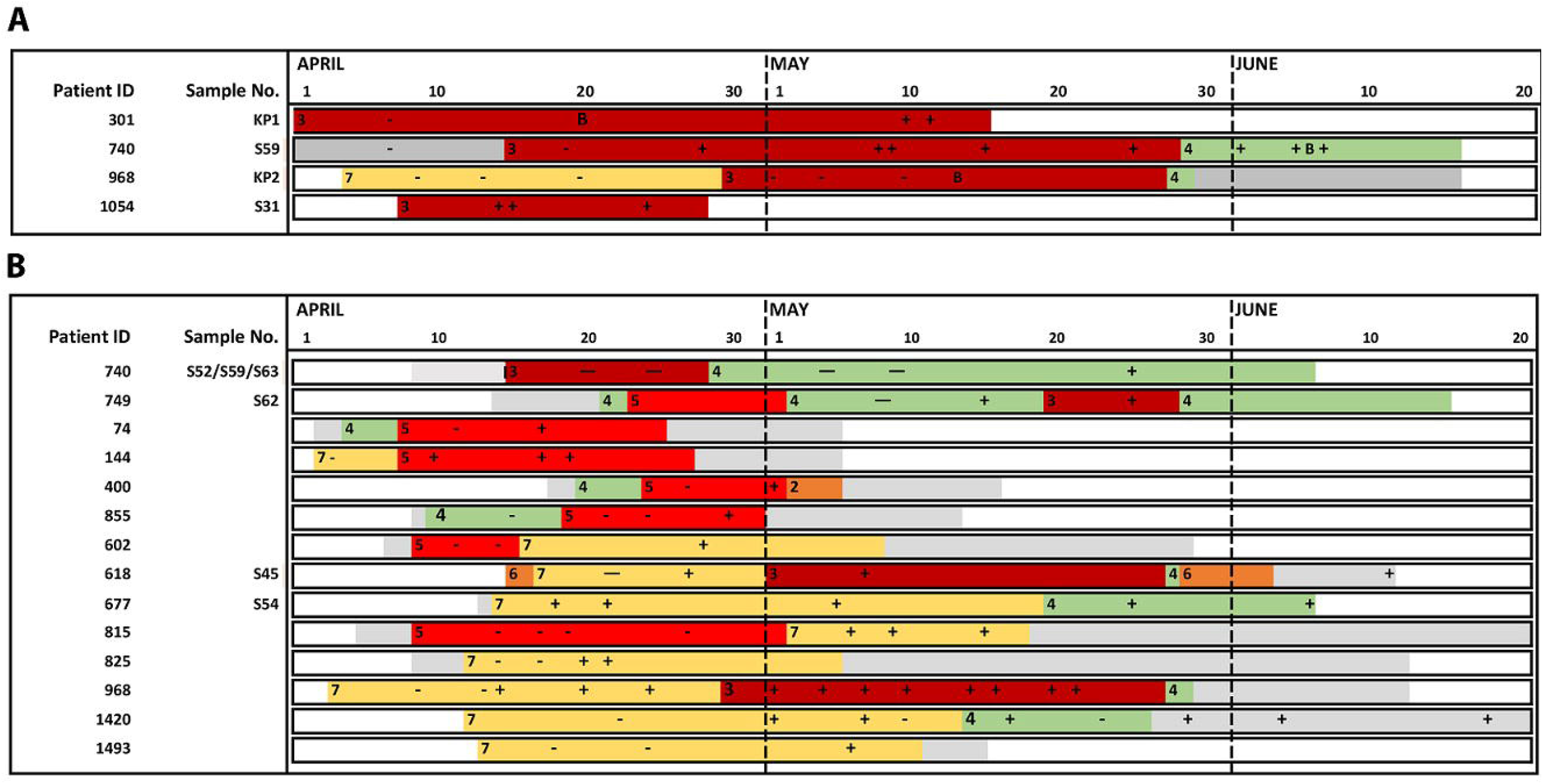
Identification of MDR *K. pneumoniae* and *C. striatum* outbreaks across the ICU network based on combined epidemiological and CMg analysis. Overlapping ward stays for patients involved in putative outbreaks of A) MDR *K. pneumoniae* and B) *C. striatum*. Each row represents a unique patient. Patients are ordered by ward of first positive (ascending) and then by patient ID (ascending). Horizontal axis shows ward stays from April 1st to Jun 20th. Non ITU wards are colored in grey. ITU wards are labelled 1-10 represented by a unique colour. Periods outside the hospital are represented in white. MDR-*K. pneumoniae* or *C. striatum p*ositive and negative respiratory samples are marked as (+) or (-) respectively. Patients with a CMg-aligned sequence have an S number (respiratory sample) or KP number (blood culture) adjacent to their identification number on the left of each bar. CMg was performed on MDR-*K. pneumoniae* in respiratory samples from patients 1054 and 301 and blood stream infection isolates on patients 301 and 968 retrieved from the routine diagnostic laboratory (timepoint marked as B). Possible chain of transmission is from top to bottom. No sequenced patient could link 968 to 1517 or 618 to 740 and so were assumed to be due to cryptic transmission via other non-sequenced patients. CMg was performed on *C. striatum* in respiratory samples from patient 618, 677, 740 and 749. All other patients were linked by epidemiology only

### Klebsiella aerogenes

Consensus sequence generated using *K. aerogenes* reads from S1, S52 and S62 identified 49,007 SNPs from 4,647,134 bases in S1 and S52, (S62 was excluded due to 1.5% genome coverage). S1 and S52 were only 98.94% identical and differed in the allele leuS (14 vs 29) indicating they were not part of an outbreak.

### Corynebacterium striatum

Analysis of consensus sequence using *C. striatum* reads from 5/6 samples (S45, S52,S54, S59 and S63) from 4 patients showed 71,339 of 2,758,551 bases present in all consensus sequences with a maximum 4 SNPs - (S62 (patient 749) was excluded due to 3.2% genome coverage). Reviewing all positions where there was a base in all samples, the maximum distance was 157 SNPs from 1,486,708 bases (99.99% identity) implying they were part of an outbreak (supplementary Table 5A). Epidemiological analysis of all 18 patients with *C. striatum* identified overlapping ward-stays for 14/18 patients across three ICUs, with genome sequence data implicating an extensive outbreak associated with patient movement between ICUs (Fig.1B).

## Discussion

This study illustrates the significant challenge facing COVID-19 ICUs with high rates of secondary infection and antimicrobial resistance in a setting that had prior sustained successful antimicrobial stewardship and infection control programmes (28, 29). There were particularly high rates of *Klebsiella spp*. infection with of respiratory 74% and 79% of BSI *Klebsiella* isolates having intrinsic or acquired resistance to first-line antibiotics (piperacillin-tazobactam). We show how a single respiratory CMg test provides pathogen identification and accurate AMR-prediction within an eight-hour laboratory workflow, and SNP-typing data the following day. This demonstrates for laboratories, intensivists, pharmacists and infection control teams the full benefit of CMg, with particular relevance to ICU settings that have unpredictable epidemiology and high levels of AMR such as, is being seen with COVID-19. Demonstrating these benefits is necessary to support the introduction of CMg into predominantly culture-based microbiology laboratories, and for the multidisciplinary team to change their clinical practice to accommodate rapid comprehensive information on ICU-pathogens. Previous studies have given examples of how CMg can diagnose respiratory infection (20, 23, 30-35), predict AMR (18, 36, 37) and provide genotyping data (37-39), but here for the first time, all these outputs are combined in a single test demonstrating the impact CMg would have when applied in a challenging real-world setting.

CMg was 93% sensitive and 81% specific for bacterial and fungal detection, consistent with previous estimates (20, 40-41). Discrepancies were mostly within polymicrobial samples with scanty growth of undetected pathogens (2 false negatives), which makes their clinical significance questionable, or where culture potentially missed the pathogen (3 false positives). Culture is a recognised imperfect gold standard, meaning specificity is likely to have been underestimated. These discordant results are not of major concern, as CMg thresholds can be refined further, allowing future translation of CMg into service evaluation.

We assessed the impact of 2-hour CMg AMR results against the updated ICU antimicrobial treatment guideline for COVID-19 patients that recommended piperacillin-tazobactam as first-line therapy, which is common practice in the UK (43, 44). CMg accurately detected acquired β-lactam genes conferring phenotypic resistance to recommended antibiotics for the main respiratory pathogens, particularly *Enterobacteriales*. Speciation or ESBL-detection would inform meropenem therapy in 33% and 19% of cases respectively rather than piperacillin-tazobactam. This is significant given meropenem improves survival compared with piperacillin-tazobactam for patients with ESBL-*Enterobacterales* infection (42). Conversely in 43% of cases combined speciation and absence of detected β-lactamases would inform co-amoxiclav therapy, aiding antimicrobial stewardship. Thus CMg-results for the 21 samples would not inform piperacillin-tazobactam use in any case. We did not demonstrate ability to detect MRSA or carbapenemase-producing *Enterobacterales* as they were absent from our cohort, however this is expected to be feasible using the same methods. CMg also detected an ESBL in one *K. pneumoniae* sample that was not phenotypically confirmed; whilst this lead to unnecessary escalation to meropenem the antibiotic suggested by CMg is still effective against the organism. Predicting AMR in *P. aeruginosa* and some other non-fermentors using CMg has not been demonstrated here, with further work required to delineate genotypic correlates with phenotypic resistance to allow same-day resistance prediction in these organisms. Nevertheless, this study illustrates the huge impact routine CMg could have on treating ICU infections, particularly where there are high or changing rates of resistance.

CMg also demonstrated potential to help with diagnosing IPA. It detected all culture positive samples and was 100% concordant with targeted qPCR, whereas half the patients with a positive GM result were not confirmed by the other three testing methodologies. Diagnosing secondary-IPA is difficult with severe viral infections (5, 45, 46) and particularly COVID-19 patients, who commonly fulfil all radiological, clinical and host diagnostic criteria (47)(24). IPA in COVID-19 patients, was uncommon in our study (about 2%) as in other London centres (26, 48). Albeit, the single small focus of IPA in only one *post-mortem* reported here and elsewhere (49-51), suggests COVID-19-related IPA may not be as clinically-significant as with influenza. Thus taken together, CMg shows potential as a rapid diagnostic for IPA, although further studies in other settings are required (52).

Finally, 24 hour CMg data identified outbreaks that help explain the epidemiology of secondary infection on COVID-19 ICUs. It identified an MDR-*K. pneumoniae* ST307 outbreak which is a particular concern given its resistance profile and extensive international spread (54, 55). Hospital transmission may therefore help explain the high prevalence of *Klebsiella spp*. reported here and potentially elsewhere (44, 53). CMg also identified an MDR-*C. striatum* outbreak potentially involving 14 patients. The clinical significance of detecting *C. striatum* in respiratory specimens is unclear although MDR-*C. striatum* outbreaks (as noted here) are also reported elsewhere (56-58). These findings highlight again the benefit of unbiased pathogen detection using CMg in revealing hidden outbreaks.

Further work is required to implement CMg into routine service, particularly automation of sample preparation to accommodate routine testing. Resistance prediction and SNP typing was also not possible in two samples due to low pathogen quantity present in samples. Further quality controls are also required for laboratory accreditation, and development of the bioinformatics tools to clinical standard for real-time analysis and generation of clinical reports. Finally, intensivists would need to change prescribing practice in response to CMg results, which may be challenging when advised to use narrower-spectrum antibiotics or withhold antibiotics when pathogens are not detected.

In summary, this study demonstrates how CMg testing identifies bacterial and fungal infections, AMR and hospital transmission events in a single rapid test. CMg could significantly improve the management of infection and transmission on ICU, with immediate potential benefit for COVID-19 patient cohorts that have heightened risk of secondary infection with MDR-pathogens (59). Early targeted therapy presents a clear opportunity to improve patient outcomes and antimicrobial stewardship. Further clinical evaluation of an ICU CMg service is our priority for the next wave of the COVID-19 pandemic.

## Methods

### Clinical setting and data collection

Clinical, microbiological and ward location data were collected by the primary care team from all patients with a documented SARS-CoV-2 RT-PCR positive test admitted to the 3 pre-existing and 4 newly-opened COVID-19 ICUs. 2 pre-existing ICUs doubled bed-capacity and all healthcare staff used additional personal protective equipment according to PHE guidelines. Updated ICU empiric antimicrobial guidelines recommended 3 days co-amoxiclav on admission, piperacillin-tazobactam for first suspected acquired respiratory infection and meropenem for subsequent infections or where resistance was suspected.

### Sample selection and analysis

Aliquots of surplus clinical respiratory samples, submitted by intensivists each day and selected by the infectious diseases consult team based on the patient having a high probability of secondary infection, were retrieved after routine processing. Samples were anonymised prior to submission to the research team. The clinical care team collected relevant clinical and laboratory data to create an anonymised dataset given to the research team who had no access to patient identifiable data at any time. The clinical team was not aware of the CMg results while caring for the patients. The full process for sample collection, nanopore sequencing, data linkage and anonymization was approved by a research ethical committee (North West Preston REC: reference 18/NW/0584).

### Routine microbiological processes

Respiratory samples were processed according to standard laboratory practice in an ISO15189 accredited laboratory (supplementary methods). Galactomannan (GM) antigen detection was performed by Mycology Reference Laboratory, Bristol using the Platelia™ Aspergillus Antigen kit (BIO-RAD – 62794). Definitions of respiratory pathogens were based on standard criteria as previously done by other studies (20, 40) - non-respiratory pathogens identified are listed in Supplementary Table 6.

### *A. fumigatus* qPCR assay

Probe-based qPCR assay was performed to amplify and detect *A. fumigatus* DNA using the QuantStudio 7 Flex (Applied Biosystems). Reagents and reactions were set up as previously described in (20) and (27) respectively.

### Nanopore metagenomic sequencing

Host DNA depletion, microbial DNA extraction and sequencing was performed using previously published methods (20) with minor modifications presented in detail in supplementary methods. Samples were batched for CMg-sequencing (6 samples per run). Library preparation was performed using the Rapid PCR Barcoding Kit (ONT) with a 6 min extension time as previously described (20). Library was loaded onto nanopore flow cells (R9.4.1) with sequencing performed on the GridION platform. ONT MinKNOW software (version 3.6.5) acquired raw sequence data with live basecalling by ONT Guppy (version 3.2.10). Sequencing was run for 24 hours with the first 2 hour data used for pathogen identification by WIMP analysis. Human reads were discarded by alignment with genome reference (GCA_000001405.15, assembly GRCh38.p13 version) and non-human reads were exported and used for pathogen identification and AMR gene detection as previously described (20).

### Pathogen identification and acquired resistance gene prediction

EPI2ME Antimicrobial Resistance pipeline (ONT, version v2020.2.10-3247478) was used for bacterial and fungal pathogen identification as previously described (20). Potential bacterial pathogen(s) were reported if ≥1% of total microbial reads and centrifuge score ≥2504. *Aspergillus spp*. were reported if ≥10 reads and a centrifuge score ≥2504. To remove possible contamination and barcode cross-talk, 0.1% of total pathogenic reads of any pathogens with >10,000 classified cumulative microbial reads were removed from all channels. Any remaining pathogens in the negative control (>5 classified reads) were considered contaminants and were removed from all the channels.

Acquired resistance genes were detected from 2 hours of sequencing with Scagaire (https://github.com/quadram-institute-bioscience/scagaire) using Abricate analysis as an input (https://github.com/tseemann/abricate). Clinically-relevant gene alignments with >90% coverage were removed and only resistance genes with >1 gene alignment were reported to remove possible bioinformatics errors (suppl. Table 3). Analysis for acquired genotypic resistance was performed for β-lactamases that impact on guideline-directed antibiotic choices (supplementary methods).

### Nanopore sequencing of *K. pneumoniae* BSI-isolates

DNA was extracted from stored *K. pneumoniae* BSI isolates by bead beating for 4m/s for 40s seconds using Matrix E beads (MP Biomedicals™) on MP Biomedicals™ FastPrep-24™ 5G Instrument (MP Biomedicals™) - see supplementary methods. Extracted DNA was washed with 0.5X of Agencourt AMPure XP beads (Beckman Coulter-A63881) and prepared for nanopore sequencing using the Native barcoding genomic DNA (ONT - EXP-NBD114 and SQK-LSK109 kits). Isolates were sequenced on a GridION for 48 hours, following manufacturer’s instructions.

### *Klebsiella spp*. and *C. striatum* SNP analysis

Representative complete reference genomes for each species were downloaded from RefSeq to generate consensus sequences (63) for *K. pneumoniae* reads from 8 patients (8 samples), *C. striatum* reads in 6 samples (4 patients) and *K. aerogenes* reads from 4 samples (3 patients) (supplementary methods). SNP-sites (v2.5.1) (66) was used to identify SNPs between each sample and SNP distances were calculated using SNP-dists (v0.7.0) (https://github.com/tseemann/snp-dists). Multi-locus sequence typing was performed using mlst (v2.19.0) (https://github.com/tseemann/mlst).

### Data availability

Sequence data presented in this study can be accessed on the European Nucleotide Archive (ENA) - study accession number PRJEB41184.

## Data Availability

https://www.ebi.ac.uk/ena/browser/view/PRJEB41184

## Acknowledgments

This research was funded/supported by the National Institute for Health Research (NIHR) Biomedical Research Centre based at Guy’s and St Thomas’ National Health Service (NHS) Foundation Trust and King’s College London, the programme of Infection and Immunity (RJ112/N027) J.D.E and T.C. J.O.G was supported by the Biotechnology and Biological Sciences Research Council (BBSRC) Institute Strategic Programme Microbes in the Food Chain BB/R012504/1 and its constituent projects BBS/E/F/000PR10348, BBS/E/F/000PR10349, BBS/E/F/000PR10351, and BBS/E/F/000PR10352 and Innovate UK-China AMR grant TS/S00887X/1. A.J.P. was supported by the Quadram Institute Bioscience BBSRC funded Core Capability Grant (project number BB/CCG1860/1). We thank Dr. Vivek Sekhwat (Speciality Trainee, Histopathology) and Ms Lara Iredale (Senior Anatomical Pathology Technologist) for contributing to the supplementary histopathology Figure 1.

## Author contributions

The study was designed by J.D.E, J.O.G., and T.C. Clinical data were collected by J.D.E, L.B.S, T.G.S.M., C.I.S.M., C.M., A.G., U.M., Laboratory work and data analysis were performed by T.C., A.AM., L.B.S and A.J.P. Clinical samples were collected and analysed by A.AM. and L.B. S.G. and D.R. All authors contributed to the write-up and review of the manuscript.

