## Supplementary material for "Application of respiratory metagenomics for COVID-19 patients on the intensive care unit to inform appropriate initial antimicrobial treatment and rapid detection of nosocomial transmission"

**Supplementary Figures:**

**Supplementary Figure 1. Post mortem histological analysis of focal invasive pulmonary aspergillosis (IPA)**. A. Focus of invasive aspergillosis in the left lower lobe involving a large vessel with surrounding lung showing diffuse alveolar damage (approximately 1cm x 1cm) (Haematoxylin & Eosin x 12). B. This was a single focus identified from ten 3 x 2cm sections of lung. B. Focal invasive aspergillosis was identified microscopically at post-mortem (Grocott silver stain x 100).

**Tables:**

**Supplementary Table 1**. Proportion of patients with different *Klebsiella spp.* in respiratory and blood cultures and their acquired resistance phenotypes impacting on guideline recommendations.

**Supplementary Table 2A.** Sequencing metadata for all respiratory samples processed with clinical metagenomics.

**Supplementary Table 2B**. Negative controls run with each batch of samples sequenced.

**Supplementary Table 3**. Phenotypic resistance reported by culture and resistance genes reported by clinical metagenomics in all culture-positive samples after 2 hours of sequencing.

**Supplementary Table 4.** Microbiology, PCR and clinical metagenomics results for all samples processed in this study for the identification of *Aspergillus fumigatus.*

**Supplementary Table 5.** *Klebsiella pneumoniae* and *Corynebacterium striatum* alignment for outbreak analysis. (A) Number of SNPs between each sample of *Corynebacterium striatum*. (B) Number of SNPs between each sample of *K. pneumoniae*, (C) the 7 predicted gene multi-locus sequence types for each sample against the *K. pneumoniae* database (numbers in each gene column refer to the allele - a ~ indicates a full length allele similar to the given allele but less than 100% identity) and (D) number of SNPs analysis in between the two identical CMg samples and two epidemiologically linked *K. pneumoniae* isolates.

**Supplementary Table 6.** All organisms identified in all respiratory samples processed with clinical metagenomics (above pre-defined thresholds[a]).

**Supplementary Table 1**. Proportion of patients with different *Klebsiella spp.* in respiratory and blood cultures and their acquired resistance phenotypes impacting on guideline recommendations

| ***Klebsiella spp*. identified** | **Number in respiratory tract (%)** | **Acquired β-lactam Resistance** | **Acquired resistance phenotypes (number)** | **Number in blood (%[f])** |
|---|---|---|---|---|
| *K. pneumoniae* | 51[a] | 32 (63%) | ESBL + AGR (13) | 3 (23%) |
| | | | Other: non-ESBL + AGS [d] (16) | 6 (30%) |
| | | | Other: AGS [e] (4) | 0 |
| | | | None (28) | 3 (11%) |
| *K. aerogenes* | 37[a] | 0[c] | N/A | 5 (14%) |
| Other *Klebsiella spp*[b] | 6 | 1 | N/A | 2 [g] (33%) |

[a] Includes 6 patients with *K. pneumoniae* and *K. aerogenes* in respiratory specimens at different times. Where total numbers in different columns or rows differ this is due to co-infection with different *Klebsiella spp.* or their resistance phenotypes at some time during ICU stay.

[b] 5 *K. oxytoca* and 1 *K. variicola*

[c] *K. aerogenes* intrinsic resistance to co-amoxiclav and pipercillin-tazobactam

[d] 14 resistant to co-amoxiclav and piperacillin-tazobactam and 6 others

[e] Co-amoxiclav susceptible but with resistance to second or third generation cephalosporins)

[f] % = Number of patients with Klebsiella BSI/number with Klebsiella in respiratory tract.

[g] 1 *K. oxytoca* with no acquired resistance and 1 *K. variicola* with resistance to co-amoxiclav and pipercillin-tazobactam

AGR = aminoglycoside resistance. AGS =aminoglycoside susceptible

**Supplementary Table 2A.** Sequencing metadata for all respiratory samples processed with clinical metagenomics.

| Sample ID | Human reads | Microbial classified reads | Unclassified reads | Microbial reads | Number of raw reads from 2hrs | Metagenomics output | Classified reads of reported pathogens after chosen thresholds[a] |
|---|---|---|---|---|---|---|---|
| S1[N3] | 2,805 | 144,220 | 975 | 145195 | 148,000 | *K. aerogenes* | 138,626 |
| S5[N3] | 16 | 0 | 0 | 0 | 16 | Negative | 0 |
| S8[N5] | 736 | 579 | 820 | 1,399 | 2135 | *A. fumigatus*<br>*K. oxytoca* | 77<br>44 |
| S10[N5] | 4,118 | 74,891 | 459 | 75,350 | 79,468 | *K. pneumoniae* | 69,029 |
| S11[N5] | 39,675 | 28,473 | 7,195 | 35,668 | 68,148 | *K. pneumoniae* | 16,828 |
| S14[N6] | 3,795 | 195 | 10 | 205 | 4000 | Negative | 0 |
| S16[N6] | 8,308 | 42,269 | 1,423 | 43,692 | 52,000 | *S. aureus* | 1,768 |
| S17[N7] | 6,095 | 1790 | 115 | 1,905 | 8000 | *P. aeruginosa* | 1,457 |
| S18[N6] | 17,224 | 19,478 | 3,298 | 22,776 | 40,000 | Negative | 0 |
| S19[N6] | 37,629 | 6,866 | 3,505 | 10,371 | 48,000 | Negative | 0 |
| S20[N7] | 21,292 | 38,361 | 347 | 38,708 | 60,000 | *S. aureus* | 36,281 |
| S21[N6] | 134 | 75,112 | 754 | 75,866 | 76000 | *E. cloacae* | 62,314 |
| S25[N15] | 62,587 | 1325 | 88 | 1,413 | 64,000 | Negative | 0 |
| S27[N11] | 11,214 | 4,238 | 548 | 4,786 | 16,000 | Negative | 0 |
| S28[N11] | 87,316 | 43,293 | 1,391 | 44,684 | 132000 | *S. aureus*<br>*A. fumigatus* | 3,165<br>2,649 |
| S29[N10] | 1108 | 439 | 320 | 759 | 1867 | Negative | 0 |
| S30[N10] | 200 | 91 | 136 | 227 | 307 | Negative | 0 |
| S31[N10] | 200 | 31,233 | 567 | 31,800 | 32,000 | *K. pneumoniae* | 28,056 |
| S33[N10] | 16,179 | 981 | 195 | 1,176 | 17,355 | Negative | 0 |

| | | | | | | | |
|---|---|---|---|---|---|---|---|
| S34[N10] | 27,481 | 76,803 | 5,993 | 82,796 | 110,277 | *K. pneumoniae* | 38,758 |
| S35[N10] | 18 | 465 | 388 | 853 | 871 | *A. baumanni* | 99 |
| S36[N8] | 119 | 117,999 | 1,882 | 119,881 | 120,000 | *S. aureus* | 109,767 |
| S37[N8] | 10,605 | 31,895 | 13,500 | 45,395 | 56,000 | *M morgannii*<br>*K. pneumoniae*<br>*P. mirabilis* | 28,300<br>876<br>397 |
| S39[N8] | 1,749 | 25804 | 447 | 26251 | 28,000 | *C. koseri*<br>*K. pneumoniae* | 23,870<br>284 |
| S40[N11] | 78 | 1024 | 133 | 1157 | 1235 | Negative | 0 |
| S41[N8] | 933 | 24,770 | 6,297 | 31,067 | 32,000 | *S. aureus* | 1,365 |
| S42[N8] | 449 | 45474 | 6,077 | 51551 | 52,000 | *Burkholderia spp.* | 34,347 |
| S44[N13] | 2,922 | 63,370 | 1,708 | 65,078 | 68,000 | *S. marcesens*<br>*C. freundi*<br>*K. aerogenes*[b] | 53,082<br>6,082<br>173 |
| S45[N16] | 12,239 | 13,256 | 2,505 | 15,761 | 28,000 | *C. striatum*<br>*K. aerogenes*[b] | 8,665<br>104 |
| S46[N13] | 766 | 300 | 162 | 462 | 1,228 | *C. koseri* | 237 |
| S49[N13] | 33,798 | 2,070 | 132 | 2,202 | 36,000 | *K. pneumoniae* | 594 |
| S50[N13] | 5,845 | 10,005 | 150 | 10,155 | 16,000 | Negative | 0 |
| S51[N14] | 87,418 | 8,342 | 240 | 8,582 | 96,000 | *S. aureus*<br>*C. koseri* | 5,203<br>2,262 |
| S52[N7] | 49,146 | 37,875 | 979 | 38,854 | 88,000 | *K. aerogenes*<br>*C. striatum* | 5,277<br>24,347 |
| S53[N15] | 24 | 0 | 0 | 0 | 24 | Negative | 0 |
| S54[N14] | 8,985 | 2,698 | 317 | 3,015 | 12,000 | *C. striatum* | 1,758 |
| S55[N11] | 26,854 | 8,724 | 422 | 9,146 | 36,000 | *A. fumigatus* | 16 |

| | | | | | | | |
|---|---|---|---|---|---|---|---|
| S56[N11] | 3,342 | 36,278 | 380 | 36,658 | 40,000 | *P. mirabilis*<br>*A. fumigatus* | 11,323<br>79 |
| S59[N14] | 542 | 117,780 | 1,678 | 119,458 | 120,000 | *K. pneumoniae*<br>*C. striatum* | 99,186<br>8,478 |
| S61[N15] | 76 | 47,159 | 765 | 47,924 | 48,000 | *C. koseri*<br>*K. pneumoniae*<br>*P. mirabilis* | 29,797<br>14,118<br>815 |
| S62[N14] | 10,979 | 920 | 101 | 1,021 | 12,000 | *K. aerogenes*<br>*C. striatum* | 184<br>227 |
| S63[N15] | 621 | 141,567 | 5,812 | 147,379 | 148,000 | *C. striatum*<br>*K. pneumoniae* | 89,166<br>17,034 |
| S64[N16] | 3,516 | 19,652 | 4,832 | 24,484 | 28,000 | Negative | 0 |
| S65[N16] | 26,010 | 32,859 | 13,131 | 45,990 | 72,000 | Negative | 0 |

[a]Pre-defined thresholds: ≥1%microbial classified, >2503 alignment score and barcode cross-talk rule applied.
[b]Pathogens identified but were not above pre-defined thresholds

**Supplementary Table 2B**. Negative controls run with each batch of samples sequenced.

| Control number | Human reads | Microbial classified reads | Unclassified reads | Number of reads minus hg38 | Number of raw reads from 2hrs | Pathogen identified by metagenomics | Pathogen classified reads after 2hrs of sequencing |
|---|---|---|---|---|---|---|---|
| N3 | 35 | 0 | 0 | 0 | 35 | *K. aerogenes*<br>*M. catarhallis*<br>*P. aeruginosa* | 23<br>6<br>5 |
| N5 | 2 | 136 | 112 | 248 | 250 | *E. coli*<br>*K. pneumoniae* | 9<br>4 |
| N6 | 2946 | 898 | 156 | 1054 | 4,000 | *E. coli* | 76 |
| N7 | 0 | 0 | 9 | 0 | 9 | - | 0 |
| N8 | 414 | 89 | 10 | 99 | 513 | *E. coli*<br>*S. aureus* | 9<br>4 |
| N10 | 5812 | 1599 | 288 | 1887 | 7699 | *E. coli*<br>*K. pneumoniae* | 390<br>20 |
| N11 | 238 | 1234 | 114 | 1348 | 1586 | *E. coli*<br>*H. influenzae*<br>*K. pneumoniae* | 290<br>14<br>11 |
| N13 | 41,567 | 2,420 | 13 | 2,433 | 44,000 | *E. coli*<br>*C. striatum* | 63<br>7 |
| N14 | 15 | 402 | 63 | 465 | 480 | *E. coli*<br>*K. pneumoniae* | 37<br>15 |
| N15 | 56 | 80 | 8642 | 6942 | 6998 | *E. coli*<br>*C. striatum*<br>*K. pneumoniae* | 13<br>3<br>9 |
| N16 | 1248 | 480 | 0 | 480 | 1728 | - | 0 |

**Supplementary Table 3**. Phenotypic resistance reported by culture and resistance genes reported by clinical metagenomics in all culture-positive samples after 2 hours of sequencing.

| Sample number | Pathogen identified by culture | Reported phenotypic resistance[a] | AMR genes reported by metagenomic sequencing |
|---|---|---|---|
| S1 | *K. aerogenes* | Amoxicillin<br>Co-amoxiclav<br>Cefuroxime<br>Ceftazidime<br>Pireracillin/Tazobactam | ND |
| S8 | *A. fumigatus* | ND | ND |
| S10 | *K. pneumoniae* | Amoxicillin | *oqcA_1*<br>*fosA_5* |
| S11 | *K. pneumoniae* | Amoxicillin | *oqxB_1*<br>*oqcA_1*<br>*fosA5_1*<br>*fosA_6*<br>*fosA_5*<br>*fosA_5*<br>*oqcA_1* |
| S17 | *P. aeruginosa* | Sensitive | ND |
| S20 | *S. aureus* | Penicillin<br>Erythromycin<br>Fusidic acid | *fusB_1*<br>*fosD_1*<br>*aadD_1*<br>*erm(T)_2* |
| S21 | *E. cloacae* | Ampicillin | *fosA_1*<br>*fosA_7* |
| S28 | *Aspergillus* | ND | ND |
| S31 | *K. pneumoniae* | ESBL<br>Ciprofloxacin<br>Penicillins<br>Septrin<br>Amoxicillin | *aph(6)-Id_1*<br>*oqxB_1*<br>*aph(3'')-Ib_5*<br>*blaOXA-1_1*<br>*dfrA14_1*<br>*oqxA_1*<br>*fosA6_1*<br>*sul2_2*<br>*blaTEM-1B_1*<br>*blaSHV-28_1*<br>*sul2_15*<br>*blaCTX-M-103_1* |
| S34 | *K. pneumoniae* | Amoxicillin | *oqxB_1 aph(3'')-Ia_5*<br>*blaTEM-168_1*<br>*fosA_3*<br>*tet(W)_4*<br>*blaSHV-108_1* |
| S35 | *A. baumannii* | Co-amoxixlav<br>Cefuroxime | No genes |

| | | | |
|---|---|---|---|
| | | Piperacillin/Tazocin<br>Cefpodoxime<br>Cefoxitin | |
| S36 | *S. aureus* | Penicillin,<br>Erythromycin<br>Ciprofloxacin<br>Trimethoprim | *ermC_13*<br>*ermC_1*<br>*ermC_10*<br>*ermC_2*<br>*ermC_12*<br>*dfrG_1* |
| S37 | *P. mirabilis* | Sensitive | *aac(3)-Iid_1*<br>*blaOXA-1_1*<br>*dfrA17_1*<br>*aadA5_1* |
| | *M morganni* | Gentamicin<br>Cephalosporin<br>Septrin<br>Ciprofloxacin<br>Trimethoprin<br>Fosfomycin | *sul1_2*<br>*aac(3)-lld_1*<br>*blaOXA-1_1*<br>*dfrA17_1*<br>*aadA5_1*<br>*blaDHA-1_1* |
| S39 | *C. koseri* | Amoxicillin | *BlaCKO-1_1*<br>*blaMAL-1_2* |
| S42 | *B. cenocepacia* | Gentamicin<br>Amikacin | ND |
| S44 | *S. marcesens* | Amoxicillin<br>Co-amoxiclav<br>Cefuroxime<br>Cotrimoxazole<br>Piperacillin/Tazobactam<br>Ceftazidime | *aac(6')-Ic_1*<br>*blaSRT-1_1* |
| | *K. aerogenes* | Amoxicillin<br>Co-amoxiclav<br>Cefuroxime<br>Piperacillin/Tazobactam<br>Ceftazidime | ND |
| S45 | *C. striatum* | ND | ND |
| S46 | *C. koseri* | Amoxicillin | ND |
| S49 | *K. pneumoniae* | Ciprofloxacin<br>ESBL | *blaTEM-168_1*<br>*blaTEM-176_1* |
| S51 | *S. aureus* | Penicillin,<br>Erythromycin | *ermC_13*<br>*ermT_2*<br>*fusB_1*<br>*blaTEM-171_1* |
| | *C. koseri* | Amoxicillin | *blaCKO-1_1* |
| S52 | *K. aerogenes* | Penicillin<br>Cephalosporin | *aac(6')-Ib_1* |
| S54 | *C. striatum* | Penicillin<br>Ciprofloxacin<br>Clindamycin | ND |
| S56 | *P. mirabilis* | Amoxicillin<br>Septrin | *aac(6')-aph(2'')_1*<br>*blaTEM-1B_1*<br>*tet(J)_2*<br>*erm(B)_18* |
| | *A. fumigatus* | ND | ND |
| S59 | *K. pneumoniae* | Penicillin<br>Cephalosporin<br>Septrin<br>Ciprofloxacin | *aph(6)-Id_1*<br>*oqxB_1*<br>*aph(3'')-Ib_5*<br>*blaOXA-1_1*<br>*dfrA14_1*<br>*oqxA_1* |

| | | | |
|---|---|---|---|
| | | | *fosA6_1*<br>*sul2_2*<br>*blaTEM-1B_1*<br>*blaSHV-28_1*<br>*sul2_15*<br>*blaSHV-106_1*<br>*sul1_2*<br>*tet(A)_4*<br>*aac(6')-Ib_1*<br>*fosA_3*<br>*blaTEM-122_1*<br>*blaCTX-M-106_1* |
| | *C. striatum* | Ciprofloxacin<br>Penicillin<br>Tetracycline<br>Clindamycin<br>Doxycycline | ND |
| S61 | *P. mirabilis* | Sensitive | No genes |
| | *K. pneumoniae* | Amoxicillin | oqxA<br>oqxB |
| S62 | *K. aerogenes* | Amoxicillin<br>Co-Amoxiclav<br>Cefuroxime<br>Piperacillin/Tazobatam<br>Ceftazidime<br>Cefpodoxime | ND |
| | *C. striatum* | Ciprofloxacin<br>Penicillin<br>Tetracycline<br>Clindamycin<br>Doxycycline | ND |
| S63 | *K. pneumoniae* | Co-amoxiclav<br>Tazocin | sul1_2<br>aac(6')-Ib<br>fosA6<br>aph(6)-Id<br>blaSHV-27 |
| | *C. striatum* | ND | ND |

[a]Intrinsic and acquired phenotypic resistance reported by culture, ND=Not Done

**Supplementary Table 4.** Microbiology, PCR and clinical metagenomics results for all samples processed in this study for the identification of *Aspergillus fumigatus*

| Patient | Respiratory samples | | | | Galactomannan (Positive/Tested) | |
|---|---|---|---|---|---|---|
| | **Sample Number** | **Organism identified by metagenomic sequencing** | ***Aspergillus fumigatus* qPCR assay (Cq)** | ***Aspergillus* Respiratory Culture** (Positive/Tested) | **BAL > 1.0** | **Serum > 0.5** |
| 26 | S35 | Negative | >40 | Negative | | |
| | Other | Not done | Not done | 0 / 1 | 0 / 1 | 0 / 1 |
| 100 | S39 | Negative | >40 | Negative | | |
| | Other | Not done | Not done | 0 / 5 | 0 / 1 | 0 / 0 |
| 121 | S37 | Negative | >40 | Negative | | |
| | Other | Not done | Not done | 0 / 5 | 0 / 0 | 0 / 0 |
| 177 | S36 | Negative | >40 | Negative | | |
| | Other | Not done | Not done | 0 / 4 | 0 / 0 | 0 / 1 |
| 196 | S42 | Negative | >40 | Negative | | |
| | Other | Not done | Not done | 0 / 2 | 0 / 0 | 0 / 0 |
| 400 | S49 | Negative | >40 | Negative | | |
| | Other | Not done | Not done | 0 / 3 | 0 / 0 | 0 / 1 |
| 408 | S21 | Negative | >40 | Negative | | |
| | Other | Not done | Not done | 0 / 3 | 0 / 0 | 0 / 0 |
| 441 | S51 | Negative | >40 | Negative | | |
| | S20 | Negative | >40 | Negative | | |
| | Other | Not done | Not done | 0 / 4 | 0 / 0 | 0 / 1 |
| 550 | S10 | Negative | >40 | Negative | | |
| | Other | Not done | Not done | 0 / 7 | 0 / 0 | 0 / 2 |
| 563 | S28 | Positive | 31 | Positive | | |
| | Other | Not done | Not done | 3 / 6 | 0 / 1 | 0 / 0 |
| 613 | S18 | Negative | >40 | Negative | | |
| | Other | Not done | Not done | 2 / 2 | 1 / 1 | 0 / 0 |
| 618 | S45 | Negative | >40 | Negative | | |
| | Other | Not done | Not done | 0 / 8 | 0 / 0 | 0 / 0 |
| 677 | S63 | Negative | >40 | Negative | | |
| | S54 | Negative | >40 | Negative | | |
| | S52 | Negative | >40 | Negative | | |
| | Other | Not done | Not done | 0 / 8 | 2 / 2 | 0 / 5 |
| 727 | S53 | Negative | >40 | Negative | | |
| | Other | Not done | Not done | 0 / 0 | 0 / 0 | 0 / 0 |
| 740 | S59 | Negative | >40 | Negative | | |
| | S30 | Negative | >40 | Negative | | |
| | Other | Not done | Not done | 0 / 16 | 1 / 4 | 1 / 2 |

| | | | | | | |
|---|---|---|---|---|---|---|
| 749 | S62 | Negative | >40 | Negative | | |
| | S40 | Negative | >40 | Negative | | |
| | Other | Not done | Not done | 0 / 8 | 0 / 1 | 0 / 1 |
| 815 | S46 | Negative | >40 | Negative | | |
| | S25 | Negative | >40 | Negative | | |
| | Other | Not done | Not done | 0 / 5 | 0 / 2 | 0 / 2 |
| 855 | S41 | Negative | >40 | Negative | | |
| | Other | Not done | Not done | 0 / 6 | 0 / 0 | 0 / 0 |
| 872 | S61 | Negative | >40 | Negative | | |
| | S11 | Negative | >40 | Negative | | |
| | Other | Not done | Not done | 0 / 4 | 0 / 1 | 0 / 3 |
| 1033 | S8 | Positive | 33 | Positive | | |
| | Other | Not done | Not done | 0 / 0 | 1 / 1 | 1 / 1 |
| 1036 | S5 | Negative | >40 | Negative | | |
| | Other | Not done | Not done | 0 / 2 | 0 / 0 | 0 / 1 |
| 1054 | S31 | Negative | >40 | Negative | | |
| | Other | Not done | Not done | 0 / 3 | 0 / 0 | 0 / 0 |
| 1065 | S19 | Negative | >40 | Negative | | |
| | S16 | Negative | >40 | Negative | | |
| | Other | Not done | Not done | 0 / 3 | 0 / 1 | 0 / 0 |
| 1069 | S17 | Negative | >40 | Negative | | |
| | Other | Not done | Not done | 0 / 3 | 0 / 0 | 0 / 1 |
| 1082 | S14 | Negative | >40 | Negative | | |
| | Other | Not done | Not done | 0 / 1 | 0 / 0 | 0 / 0 |
| 1092 | S27 | Negative | >40 | Negative | | |
| | Other | Not done | Not done | 0 / 5 | 0 / 2 | 0 / 2 |
| 1262 | S29 | Negative | >40 | Negative | | |
| | Other | Not done | Not done | 0 / 3 | 0 / 1 | 0 / 1 |
| 1292 | S44 | Negative | >40 | Negative | | |
| | Other | Not done | Not done | 0 / 5 | 0 / 3 | 0 / 1 |
| 1346 | S56 | Positive | 32 | Positive | | |
| | Other | Not done | Not done | 0 / 3 | 1 / 1 | 0 / 2 |
| 1440 | S33 | Negative | >40 | Negative | | |
| | Other | Not done | Not done | 0 / 4 | 1 / 2 | 0 / 1 |
| 1457 | S65 | Negative | >40 | Negative | | |
| | S64 | Negative | >40 | Negative | | |
| | Other | Not done | Not done | 0 / 9 | 2 / 2 | 0 / 2 |
| 1503 | S1 | Negative | >40 | Negative | | |
| | Other | Not done | Not done | 0 / 5 | 0 / 0 | 0 / 1 |
| 1512 | S34 | Negative | >40 | Negative | | |
| | Other | Not done | Not done | 0 / 3 | 0 / 1 | 0 / 2 |
| 1583 | S55 | Positive | 31 | Negative | | |
| | Other | Not done | Not done | 4 / 5 | 0 / 0 | 1 / 1 |

**Supplementary Table 5.** *Klebsiella pneumoniae* and *Corynebacterium striatum* alignment for outbreak analysis**.** (A) Number of SNPs between each sample of *Corynebacterium striatum.* (B) Number of SNPs between each sample of *K. pneumoniae,* (C) the 7 predicted gene multi-locus sequence types for each sample against the *K. pneumoniae* database (numbers in each gene column refer to the allele - a ~ indicates a full length allele similar to the given allele but less than 100% identity) and (D) number of SNPs analysis in between the two identical CMg samples and two epidemiologically linked *K. pneumoniae* isolates.

**A**

| Patient ID | Sample | S45 | S54 | S59 | S52 | S63 |
|---|---|---|---|---|---|---|
| **618** | **S45** | 0 | 61 | 115 | 92 | 102 |
| **677** | **S54** | 61 | 0 | 122 | 89 | 97 |
| **740** | **S59** | 115 | 122 | 0 | 157 | 157 |
| | **S52** | 92 | 89 | 157 | 0 | 30 |
| | **S63** | 102 | 97 | 157 | 30 | 0 |

**B**

| Patient ID | Sample | S10 | S11 | S31 | S34 | S59 | S63 | S61 |
|---|---|---|---|---|---|---|---|---|
| **550** | **S10** | 0 | 11273 | 11158 | 11322 | 11161 | 12162 | 26023 |
| **872** | **S11** | 11273 | 0 | 11216 | 11512 | 11219 | 11889 | 26148 |
| **1054** | **S31** | 11158 | 11216 | 0 | 11255 | 3 | 12081 | 25874 |
| **1512** | **S34** | 11322 | 11512 | 11255 | 0 | 11258 | 12525 | 25997 |
| **740** | **S59** | 11161 | 11219 | 3 | 11258 | 0 | 12084 | 25877 |
| **677** | **S63** | 12162 | 11889 | 12081 | 12525 | 12084 | 0 | 26929 |
| **872** | **S61** | 26023 | 26148 | 25874 | 25997 | 25877 | 26929 | 0 |

**C**

| Patient ID | Sample | ST | *gapA* | *infB* | *mdh* | *pgi* | *phoE* | *rpoB* | *tonB* |
|---|---|---|---|---|---|---|---|---|---|
| **550** | **S10** | - | 9 | ~4 | ~2 | 1 | 1 | 1 | ~27 |
| **872** | **S11** | 187 | 23 | 31 | 2 | 1 | 9 | 4 | 23 |
| **1054** | **S31** | - | ~4 | 1 | 2 | 52 | 1 | 1 | 7 |
| **1512** | **S34** | 33 | 2 | 3 | 5 | 1 | 12 | 4 | 9 |
| **400** | **S49** | - | | | | | | | |
| **740** | **S59** | 307 | 4 | 1 | 2 | 52 | 1 | 1 | 7 |
| **872** | **S61** | - | | | | | | | |
| **677** | **S63** | 661 | 4 | 3 | 1 | 36 | 9 | 10 | 14 |

**D**

| Patient ID | 301 | 968 | 1054 | 740 |
|---|---|---|---|---|
| **Sample ID** | **KP1** | **KP2** | **S31** | **S59** |
| **KP2** | 0 | 5 | 6 | 12 |
| **KP1** | 5 | 0 | 14 | 24 |
| **S31** | 6 | 14 | 0 | 55 |
| **S59** | 12 | 24 | 55 | 0 |

**Supplementary Table 6.** All organisms identified in all respiratory samples processed with clinical metagenomics (above pre-defined thresholds[a]).

| Sample ID | Organisms[b] identified above pre-defined thresholds[+] by respiratory metagenomics |
|---|---|
| S1 | N/A |
| S5 | N/A |
| S8 | *Candida albicans*<br>*Cutibacterium acnes*<br>*Cutibacterium acnes* HL096PA1<br>*Moraxella osloensis* |
| S10 | *Prevotella denticola* F0289<br>*Prevotella intermedia* |
| S11 | *Candida dubliniensis*<br>*Candida albicans* |
| S14 | N/A |
| S16 | *Candida ortholipsilosis*<br>*S. epidermidis*<br>*S. haemolyticus,*<br>*Lactobacillus paracasei*<br>*Lactobacillus casei* |
| S17 | N/A |
| S18 | *Candida glabrata*<br>*Candida dublinensis*<br>*Tannerella forsythia*<br>*Olsenella uli* |
| S19 | *C. orthopsilosis* |
| S20 | N/A |
| S21 | *Candida albicans*<br>*Candida glabrata*<br>*Enterobacter hormaechei subsp. oharae,*<br>*Enterobacter hormaechei subsp. steigerwaltii* |
| S25 | N/A |
| S27 | *Cutibacterium acnes* HL096PA1<br>*Cutibacterium acnes* KPA171202<br>*Escherichia coli 'BL21-Gold(DE3)pLysS AG'*<br>Arthrobacter sp. *IHBB 11108*<br>*Bacillus subtilis* BEST7003<br>*Jonesia denitrificans* DSM 20603<br>*Moraxella osloensis* |
| S28 | *Candida albicans*<br>*E. faecium*<br>*Staphylococcus epidermidis*<br>*Staphylococcus epidermidis* ATCC 12228<br>*Staphylococcus haemolyticus* JCSC1435<br>*Staphylococcus haemolyticus* |
| S29 | *Cutibacterium acnes*<br>*Bacillus subtilis* BEST7003<br>*Arthrobacter sp.* IHBB 11108<br>*Moraxella osloensis*<br>*Mucilaginibacter sp.* PAMC 26640<br>*Haemophilus parainfluenzae* T3T1 |
| S30 | N/A |
| S31 | N/A |
| S33 | *Escherichia coli 'BL21-Gold(DE3)pLysS AG'*<br>*Escherichia coli* |

| | |
|---|---|
| | *Cutibacterium acnes*<br>*Atopobium parvulum* DSM 204699<br>*Arthrobacter sp*. IHBB 11108<br>*Bacillus subtilis* BEST7003<br>*Moraxella osloensis* |
| S34 | *Candida albicans*<br>*Streptococcus oralis*<br>*Streptococcus oralis* Uo5<br>*Prevotella melaninogenica*<br>*Prevotella sp. oral taxon 299 str.* F0039 |
| S35 | *Acinetobacter pittii,*<br>*Acinetobacter johnsonii* XBB1<br>*Acinetobacter sp.* TTH0-4<br>*Acinetobacter schindleri*<br>*Acinetobacter sp*. NCu2D-2<br>Acinetobacter nosocomialis<br>Acinetobacter haemolyticus |
| S36 | N/A |
| S37 | *Neisseria sicca*<br>*Neisseria elongata subsp. glycolytica* ATCC 29315 |
| S39 | N/A |
| S40 | N/A |
| S41 | *Candida albicans*<br>*Eikenella corrodens,*<br>*Streptococcus constellatus subsp. pharyngis*<br>*Streptococcus anginosus* C238<br>*Streptococcus anginosus subsp. whileyi* MAS624<br>*Streptococcus intermedius* JTH08<br>*Staphylococcus epidermidis*<br>*Prevotella melaninogenica*<br>*Prevotella intermedia*<br>*Rothia dentocariosa* ATCC 17931 |
| S42 | *Burkholderia thailandensis*<br>*Burkholderia thailandensis* 2002721643<br>*Burkholderia pseudomallei* |
| S44 | N/A |
| S45 | *Corynebacterium simulans*<br>*Corynebacterium resistens* DSM 45100 |
| S46 | *Cutibacterium acnes* |
| S49 | *Candida albicans*<br>*Streptococcus sanguinis* SK36<br>*Bacillus subtilis* BEST7003<br>*Arthrobacter sp.* IHBB 11108<br>*Staphylococcus epidermidis*<br>*Enterococcus faecium*<br>*Cutibacterium acnes*<br>*Mucilaginibacter sp.* PAMC 26640 |
| S51 | N/A |
| S52 | *Corynebacterium simulans*<br>*Candida albicans* |
| S53 | N/A |
| S54 | *Corynebacterium simulans,*<br>*Corynebacterium resistens* DSM 45100<br>*Cutibacterium acnes*<br>*Corynebacterium aurimucosum* ATCC 700975<br>*Corynebacterium resistens* DSM 45100 |
| S55 | *Sphingomonas sp.* LK11 |

| | |
|---|---|
| S56 | *E. faecium* |
| S59 | *Corynebacterium simulans* |
| S61 | N/A |
| S62 | *Corynebacterium simulans*<br>*Arthrobacter sp.* IHBB 11108<br>*Bacillus subtilis* BEST7003<br>*Staphylococcus epidermidis*<br>*Mucilaginibacter sp*. PAMC 26640<br>*Corynebacterium kroppenstedtii* DSM 44385<br>*Cutibacterium acnes* |
| S63 | *Corynebacterium simulans*<br>*Corynebacterium aurimucosum* ATCC 700975<br>*Corynebacterium resistens* DSM 45100 |
| S64 | *Candida albicans*<br>*Prevotella melaninogenica,*<br>*Streptococcus gordonii,*<br>*Streptococcus gordonii str. challis substr.* CH1<br>*Actinomyces pacaensis*<br>*Prevotella intermedia*<br>*Fusobacterium nucleatum subsp. vincentii*<br>*Prevotella fusca* JCM 17724,<br>*Parabacteroides sp.* CT06,<br>*Porphyromonas gingivalis* |
| S65 | *Candida albicans*<br>*Prevotella melaninogenica*<br>*Atopobium parvulum* DSM 20469,<br>*Streptococcus oralis*<br>*Streptococcus oralis subsp. tigurinus*<br>*Prevotella intermedia*<br>*Fusobacterium nucleatum subsp. vincentii,*<br>*Fusobacterium nucleatum subsp. vincentii* 3_1_36A2<br>*Streptococcus gordonii*<br>*Porphyromonas gingivalis*<br>*Parabacteroides sp. CT06* |

[a]pre-defined thresholds are: ≥1%microbial classified, ≥2504 alignment score and barcode cross-talk rule. [b]Not defined as pathogenic organisms in this study
