## Supplementary methods for "Application of respiratory metagenomics for COVID-19 patients on the intensive care unit to inform appropriate initial antimicrobial treatment and rapid detection of nosocomial transmission"

### Routine microbiological processes

ICU respiratory samples were centrifuged and the pellet resuspended before streaking onto blood agar, chocolate agar, FAA (fastidious anaerobic agar) and Sabouraud agar. Sputum and Endotracheal tube (ETT) samples were directly streaked onto blood agar and chocolate agar plates. Plates were incubated for 48 hrs apart from Sabouraud agar plates that were incubated aerobically for 5 days for selective detection of *Candida* spp. and *Aspergillus* spp. Colonies were identified using MALDI-TOF (Bruker) except the *Aspergillus spp.* where microscopy was performed. Antibiotic susceptibility was performed by agar diffusion following guidelines of  the European Committee on Antimicrobial Susceptibility Testing (EUCAST) methodology (1). In this study microorganisms referred as 'Respiratory pathogens' or 'pathogens' were defined as agents causing respiratory infection based on the list used by clinical microbiology for reporting respiratory pathogens (2). Respiratory pathogens identified in this study were: *A. baumannii, A. fumigatus, B. cepacia, B. cenocepacia, C.koseri, C. striatum, E. cloacae complex, E. coli, H. influenzae, K. aerogenes, K. oxytoca, K. pneumoniae, M. catarrhalis, M.morganii, P. mirabilis, P. aeruginosa, S. marcescens, S maltophilia,* and *S. aureus.* The only fungal organisms reported as a respiratory pathogens was *A. fumigatus.* Microorganisms identified in this study (above chosen thresholds) but are not defined as respiratory pathogens are listed in Supplementary Table 6.

### Routine SARS-CoV-2 RT-PCR

200µl of clinical samples (Nose and Throat swabs or BALs) were mixed with a lysis buffer comprising Buffer ACL (contains guanidinium thiocyanate), Buffer ATL (contains sodium dodecyl sulphate), Proteinase K, MS2, EXO IPC and carrier RNA and heated at 68°C for 15 min to inactivate viruses in the sample. Following inactivation, RNA extraction of SARS-CoV-2 was performed. Automated extraction was done with the QIAsymphony SP module using the Virus Pathogen Mini kits (Qiagen - 937036) and Off-Board Lysis protocol (elution volume = 60 µl).

The RT-PCR for detection of SARS-CoV-2 was performed with the High-Plex 24 AusDiaganostic Pty Ltd, according to manufacturer's instructions using (SARS-CoV-2, Influenza and RSV 8-well, Catalogue number: 20081, Version: 08) targeting the Orf1ab and Orf8 of SARS-CoV-2. The RT-PCR reaction consisted of two steps; for the first step, a reverse transcriptase reaction is performed followed by a 15-cycle multiplexed preliminary amplification reaction. In the second step, individual real-time PCR reactions are performed using nested primers and the products from the first step. Amplification is detected by an increase in fluorescence and melt-curve analysis is performed to verify specific detection of each target.

### *Aspergillus fumigatus* qPCR assay

Probe-based qPCR assay was performed on all samples from the CMg cohort (n=43) to detect and amplify A. *fumigatus* DNA (previously described in (3)). The assay was done using the QuantStudio 7 Flex (Applied Biosystems). The master mix for each reaction consisted of 10 µl of LightCycler 480 probe master (2x), 0.4 µl of probe (final concentration 0.2 µM) and 0.5 µl each of the forward and reverse primer (final concentration 0.25 µM), 2 µl of DNA was added and nuclease free water was added to the reaction to make up volume to 20 µl. The conditions followed for all qPCR reactions were: pre-incubation at 95 °C for 15 min, amplification for 40 cycles at 94 °C for 15 s and 60 °C for 1 min.

### Galactomannan assay

Samples requested for Galactomannan (GM) antigen detection were sent to Mycology Reference Laboratory National Infection Services, PHE at Southmead Hospital, Bristol. The Platelia™ Aspergillus Antigen kit (BIO-RAD – 62794) was used according to manufacturer's instructions to detect galactomannan in sera and BALs. The assay is an one-stage immunoenzymatic sandwich microplate and uses rat EBA-2 monoclonal antibodies designed to detect *Aspergillus* GM antigens in clinical samples.

### Nanopore metagenomic sequencing

Respiratory samples were treated with sputasol (Oxoid) to liquefy samples before treatment with 1% saponin (Tokyo Chemical Industry) to induce host cell lysis and release of host DNA that was digested with DNase (Articzymes). Samples were then washed and centrifuged to pellet bacterial and fungal organisms. The pellet was re-suspended in lysis buffer (Roche UK) for bead-beating to release microbial DNA, before proteinase K treatment to digest residual proteins (Qiagen) Finally, samples were incubated at 95 °C for 30 min to kill residual organisms before DNA extraction using the Fast Pathogen 200 protocol on a MagNA Pure 24 System (Roche UK). DNA was quantified using the high sensitivity dsDNA assay kit (Thermo Fisher) on the Qubit 3.0 Fluorometer (Thermo Fisher). Fragment size and quality of metagenomic libraries were analysed using the TapeStation 4200 (Agilent Technologies) automated electrophoresis platform.

Following the second PBS wash, the pellet was re-suspended in 600 µl of bacterial lysis buffer (Roche UK - 4659180001) and total volume was transferred to a bead-beating tube (Lysis Matrix E, MP Biomedicals -116914050). Samples were bead-beaten at maximum speed (50 oscillations per second) for 1 min using a MP Biomedicals™ FastPrep-24™ 5G Instrumentin (MP Biomedicals™ - 116005500). Sample/s were centrifuged at top speed (~20,000 xg) for 1 min and ~200 µl of supernatant was carefully transferred to a fresh Eppendorf tube without disturbing the beads. 20 µl of proteinase K (>600 mAu/ml, Qiagen - 19133) was added and sample was incubated at 65°C for 5 min with shaking at 1000 RPM on an Eppendorf Thermomixer. Next samples were incubated at 95 °C for 30 min to ensure killing of any not-lysed organisms. Next, heat-killed sample was transferred to a fresh tube and DNA was extracted using the Roche MagNA Pure 24 System Fast Pathogen 200 protocol (MagNA pure 24 System Total NA Isolation Kit 1.01, Roche UK - 07658036001) on a MagNA Pure 24 System (Roche UK - 07290519001).

DNA was quantified using the high sensitivity dsDNA assay kit (Thermo Fisher - Q32851) on the Qubit 3.0 Fluorometer (Thermo Fisher - Q33226). Fragment size and quality of metagenomic libraries were analysed using the TapeStation 4200 (Agilent Technologies - G2991AA) automated electrophoresis platform with the Genomic ScreenTape (Agilent Technologies - 5067-5365) and a DNA ladder (200 to >60,000 bp, Agilent Technologies - 5067-5366).

**Pathogen identification and acquired resistance gene prediction**

Sequencing data were analysed with the EPI2ME Antimicrobial Resistance pipeline (ONT, version v2020.2.10-3247478) to identify bacterial and fungal pathogens present in the clinical samples. WIMP (What's in my Pot – tool within this pipeline) was used in this study to identify respiratory bacterial and fungal pathogens. WIMP uses 'Centrifuge' a kmer-based metagenomic classifier (4) and a pre-built database, which is based on the NCBI taxonomy and RefSeq database. (WIMP manual: https://community.nanoporetech.com/protocols/epi2me/v/mte_1014_v1_revaq_11apr2016/what-s-in-my-pot-wimp). Potential bacterial pathogen(s) were reported only if the number of reads was ≥1% of microbial reads and with a centrifuge score ≥2504. Potential fungal pathogens (i.e. *Aspergillus spp.*) were reported if ≥10 reads were classified with a centrifuge score ≥2504. A lower read count threshold was set for reporting *Aspergillus* as they are often present in low numbers in respiratory samples (all culture positive *Aspergillus* samples were reported as scanty (S) growth in this study). Additional parameters were applied to identify and remove possible contamination and barcode cross-talk: i) to eliminate barcode cross-talk, 0.1% of total pathogenic reads were removed from all channels (including process negative control) for any pathogens reported >10,000 classified microbial reads (cumulative read count observed in the multiplexed run for that pathogen); ii) any pathogens remaining in the process negative control (>5 classified reads) were considered contaminants and were removed from all the samples in the multiplex run.

Acquired resistance genes were detected from 2 hours of sequencing using Scagaire with default parameters. Scagaire utilises a bundled database containing the 40 most common-sequenced bacterial species in the RefSeq database and only reports cilinically-relevant acquired resistance genes (https://github.com/quadram-institute-bioscience/scagaire). Briefly, FASTQ files were converted into FASTA files and then analysed using Abricate (https://github.com/tseemann/abricate), with default parameters, to detect resistance genes against the ResFinder database. Then, Scagaire was used to predict medically relevant genes based on the pathogen identified by metagenomics and the Abricate output file. Clinically-relevant gene alignments with <90% coverage were removed and only resistance genes with >1 gene alignment were reported to remove any possible bioinformatics errors (suppl. Table 3).

This analysis was only carried out for specific acquired, genotypic resistances in specific Gram negative organisms and *Staphylococcus aureus*. We chose to look at acquired resistance in samples with Gram negatives present, where the Gram negatives could not be predicted harbour resistance to piperacillin-tazobactam based on pathogen identification alone. For example, in samples where *Burkholderia* spp. were identified as the sole Gram negative pathogen, were excluded. Analysis was only completed where there was concordance between identification of these organisms in both routine culture and CMg, so that resistance profiles between genotypic and routine phenotypic analysis could be compared. Samples where *Pseudomonas aeruginosa* was identified as the sole pathogen were also excluded, due to the known difficulty in predicting phenotypic resistance for this organism based on genotypic elements only (5, 6).

### Nanopore sequencing of *K. pneumoniae* BSI-isolates

Isolates of *K. pneumoniae,* previously identified by MALDI-TOF, were subcultured on blood agar and incubated for 48h at 35˚C aerobically. For bacterial DNA extraction, 4/5 colonies were selected and were mixed in 500 µl of PBS. Mixed solution was transferred into Lysing

Beads - Matrix E ((MP Biomedicals™ - 116005500) and bead-beaten for 4m/s for 40s seconds using a MP Biomedicals™ FastPrep-24™ 5G Instrument (MP Biomedicals™ - 116005500). The sample was then centrifuged for 1 min at 12000 x rpm and 100 µl of the supernatant was collected and transferred to a  clean 1.5 ml Eppendorf tube. Then, 0.5X of Agencourt AMPure XP beads (Beckman Coulter-A63881) was added, mixed and incubated for 10min at RT. The tube was placed in a magnetic rack and washed twice with 80% of ethanol before the sample was eluted in 50 µl of nuclease-free water.

### ***Klebsiella spp.*** **and** ***C. striatum*** **SNP analysis**

Representative complete reference genomes for each species were downloaded from RefSeq to generate consensus sequences (7). *K. pneumoniae* reads from 7 patients (8 samples) were aligned to the *K. pneumoniae* subsp. pneumoniae HS11286 strain. *K. aerogenes* reads from 4 samples (3 patients) were aligned to the *K. aerogenes* strain NCTC9735. *C. striatum* reads in 5 samples (4 patients) were aligned to *C. striatum* strain KC-Na-01. Reads were aligned to each matching reference genome using minimap2 (v 2.17-r941) (8). A consensus sequence was generated using bcftools (v 1.10.2) (9) and the chromosomal consensus sequences were saved as a single multi-FASTA alignment. SNP-sites (v2.5.1)(10) identified the SNPs between each sample. Multi-locus sequence typing was performed using mlst (v2.19.0) (https://github.com/tseemann/mlst). FASTQ/FASTA files were transformed using PyFASTAQ (v3.17.0)  (https://github.com/sanger-pathogens/Fastaq). SNP distances were calculated using SNP-dists (v0.7.0) (https://github.com/tseemann/snp-dists).
