## Supplementary figures and images for "Application of respiratory metagenomics for COVID-19 patients on the intensive care unit to inform appropriate initial antimicrobial treatment and rapid detection of nosocomial transmission"

### Supplemental Figure 1A

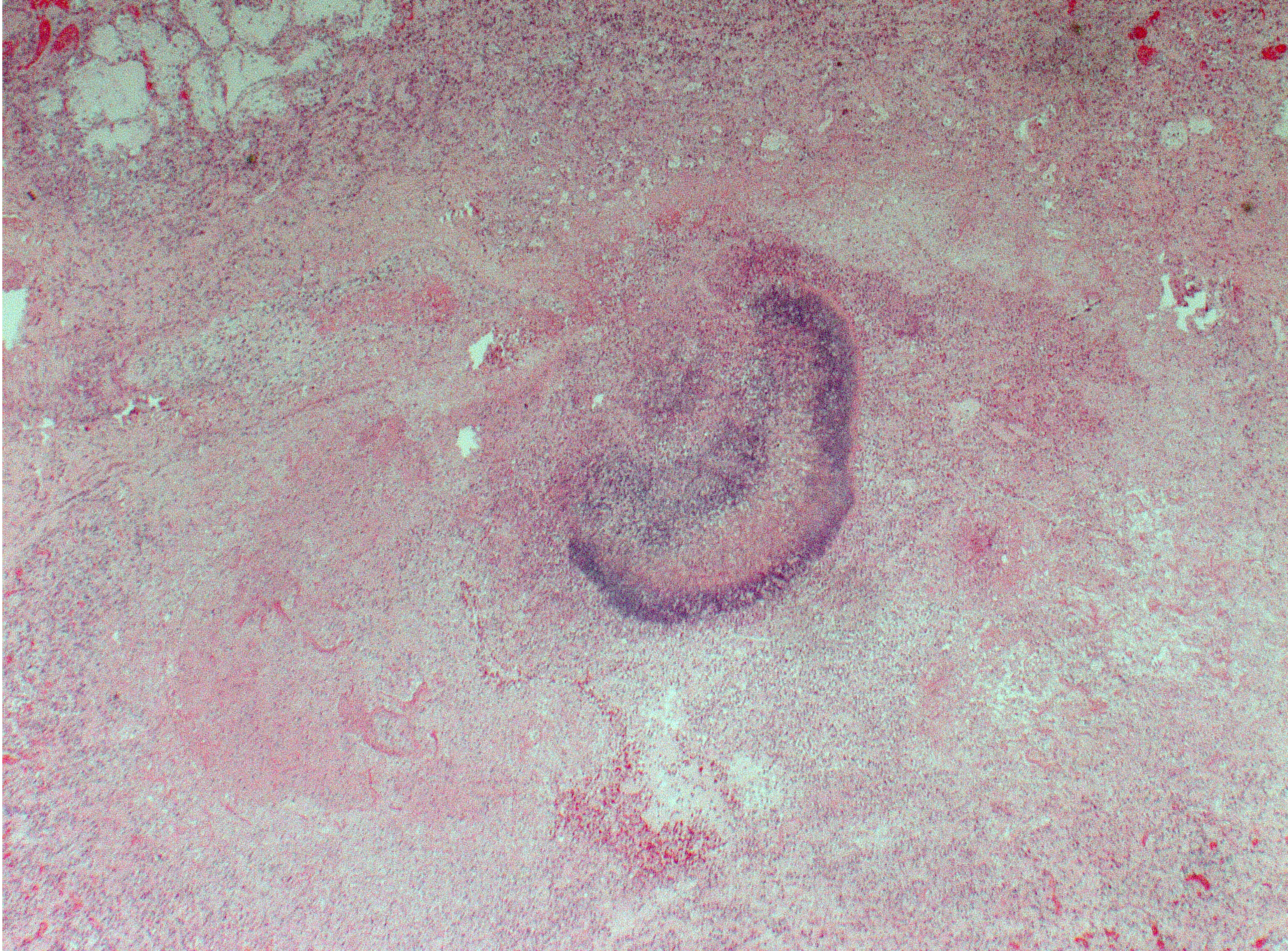

### Supplemental Figure 1B

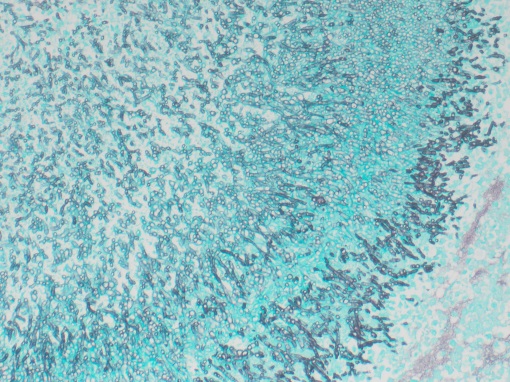
